# Clinical features, Diagnosis, and Treatment of COVID-19: A systematic review of case reports and case series

**DOI:** 10.1101/2020.03.28.20046151

**Authors:** Azin Tahvildari, Mahta Arbabi, Yeganeh Farsi, Parnian Jamshidi, Saba Hasanzadeh, Tess Moore Calcagno, Mohammad Javad Nasiri, Mehdi Mirsaeidi

## Abstract

**Objectives:** The 2019 novel coronavirus (COVID-19) has been declared a public health emergency worldwide. The objective of this systematic review was to characterize the clinical, diagnostic, and treatment characteristics of patients presenting with COVID-19.

**Methods:** We conducted a structured search using PubMed/Medline, Embase, Web of Science and the Cochrane Library to collect both case reports and case series on COVID-19 published up to February 30, 2020.

**Results:** Thirty-four articles were included analyzing a total of 99 patients with a mean age of 46.2 years. The most common presenting symptom in patients who tested positive for COVID-19 was fever, reported in up to 83% of patients from 76.4% of the analyzed studies. Other symptoms including rhinorrhea, dizziness, and chills were less frequently reported. Additionally, in studies which reported C-reactive protein (CRP) measurements (44%), a large majority of patients displayed an elevated CRP (73%). Progression to acute respiratory distress syndrome (ARDS) was the most common complication of patients testing positive for COVID-19 (33%). CT images displayed ground-glass opacification (GGO) patterns (80%) as well as bilateral lung involvement (71.0%). The most commonly used antiviral treatment modalities included, lopinavir (HIV protease inhibitor), arbidiol hydrochloride (influenza fusion inhibitor), and oseltamivir (neuraminidase inhibitor).

**Conclusions:** Development of ARDS may play a role in estimating disease progression and mortality risk. Early detection of elevations in serum CRP, combined with a clinical COVID-19 symptom presentation may be used as a surrogate marker for presence and severity of disease. There is a paucity of data surrounding the efficacy of treatments. There is currently not a well-established gold standard therapy for the treatment of diagnosed COVID-19. Further prospective investigations are necessary.

## Introduction

Late in December 2019 and early in January 2020, reports of a very progressive pneumonia-like respiratory syndrome, starting in Wuhan, China, induced global health concerns [1]. Soon after the onset of disease, it was found that the pathogen was a new member of the coronaviridae family, named SARS-COV-2 which is now called 2019-n-CoV [2]. The respiratory syndrome caused by 2019-n-CoV is called COVID-19. COVID-19 is characterized by low-grade fever, cough, dyspnea, lymphopenia, and ground-glass opacities on chest CT scan [3, 4]. COVID-19 is a highly contagious disease, probably an aerosol born one, with human to human transmission capacity which has implicated many countries all around the world [5]. In this review article, we systematically surveyed case reports and case series from many countries in the world to give a picture of the epidemiology, clinical presentations, laboratory changes, imaging findings, diagnostic criteria, treatments, outcomes, prognostic factors, and risk factors of COVID-19.

## Methods

This review conforms to the “Preferred Reporting Items for Systematic Reviews and Meta-Analyses” (PRISMA) statement [6].

### Search strategy

We carried out systematic searches of the literature in the following bibliographical databases: PubMed/Medline, Embase, Web of Science and the Cochrane Library. Search criteria included articles published in the period from January 1, 2019, to February 30, 2020, and only included articles published in English. The search terms for our review were: COVID-19, severe acute respiratory syndrome coronavirus 2, novel coronavirus, SARS-CoV-2, nCoV disease, SARS2, COVID-19, 2019-nCoV, coronavirus disease-19, coronavirus disease 2019, and 2019 novel coronavirus.

### Study Selection

Studies included in the review met the following criteria: prospective or retrospective descriptive case reports and case series of COVID-19 in the hospital setting which included diagnostic methods, clinical manifestations, laboratory features, treatment, and outcomes. Articles describing experimental approaches as well as reviews and publications without peer-review processes were excluded.

All potentially relevant articles were screened in two stages for eligibility. In the first stage, the titles and abstracts of potentially relevant articles were screened independently by two reviewers (YF, PJ). In the second stage of assessment, the full text of those abstracts which met the inclusion criteria was retrieved and independently reviewed by the same authors. Disagreements and technical uncertainties were discussed and resolved between review authors (AT, SH, MA, MJN).

### Data extraction

The extracted data included bibliographic data, patient demographics (e.g., age and gender), radiological and laboratory findings, treatment protocols, and medical consequences. Two authors (AT, SH) independently extracted the data from the selected studies. The data was jointly reconciled, and disagreements were discussed and resolved between review authors (YF, PJ, MA, MJN).

## Results

As illustrated in Figure 1, our systematic search resulted in an initial number of 1102 of potentially relevant articles, of which 346 were excluded by title and abstract evaluation. Applying the inclusion/exclusion criteria to the full-text documents, 34 articles were eligible for inclusion in the systematic review. 23 case reports and 11 case series from 9 countries were identified with a total of 99 unique cases of COVID-19. RT-PCR COVID-19 was present in 32 (94%) articles as inclusion criteria. In addition to RT-PCR, CT scan served as a diagnostic tool in 14 (41.0%) of papers. Reported comorbidities included hypertension, diabetes, cardiovascular disease, and pulmonary disease. Hypertension was investigated the most, studied in 7/34 (20%) of papers. Of the 16 COVID-19 positive patients found in the studies investigating hypertension, 8 patients were hypertensive (50%). (Table 2). Lymphopenia was reported in 10 studies which identified 12/21 (57%) of COVID-19 positive patients with lymphopenia. Additionally, in studies which reported C-reactive protein (CRP) measurements (44%), a large majority of patients displayed an elevated CRP (73%). CT imaging was used as a diagnostic and prognostic tool in 8 studies. CT images commonly displayed ground-glass opacification (GGO) patterns (80%) as well as bilateral lung involvement (71.0%). Progression to acute respiratory distress syndrome (ARDS) was the most common complication of patients testing positive for COVID-19. We found 2/34 (5.8%) reports on Acute Respiratory Distress Syndrome (ARDS), 2 of 6 (33%) investigated cases had ARDS. Morality outcomes were difficult to assess; only two studies showed mortality data in which 2/2 COVID-19 patients died. A wide range of therapeutic modalities were tried across studies, with antiviral treatments being the most used. Common antiviral treatment modalities included lopinavir (HIV protease inhibitor), arbidiol hydrochloride (influenza fusion inhibitor), and oseltamivir (neuraminidase inhibitor). In Table 3 we summarize all of the drugs used.

**Table 1.**
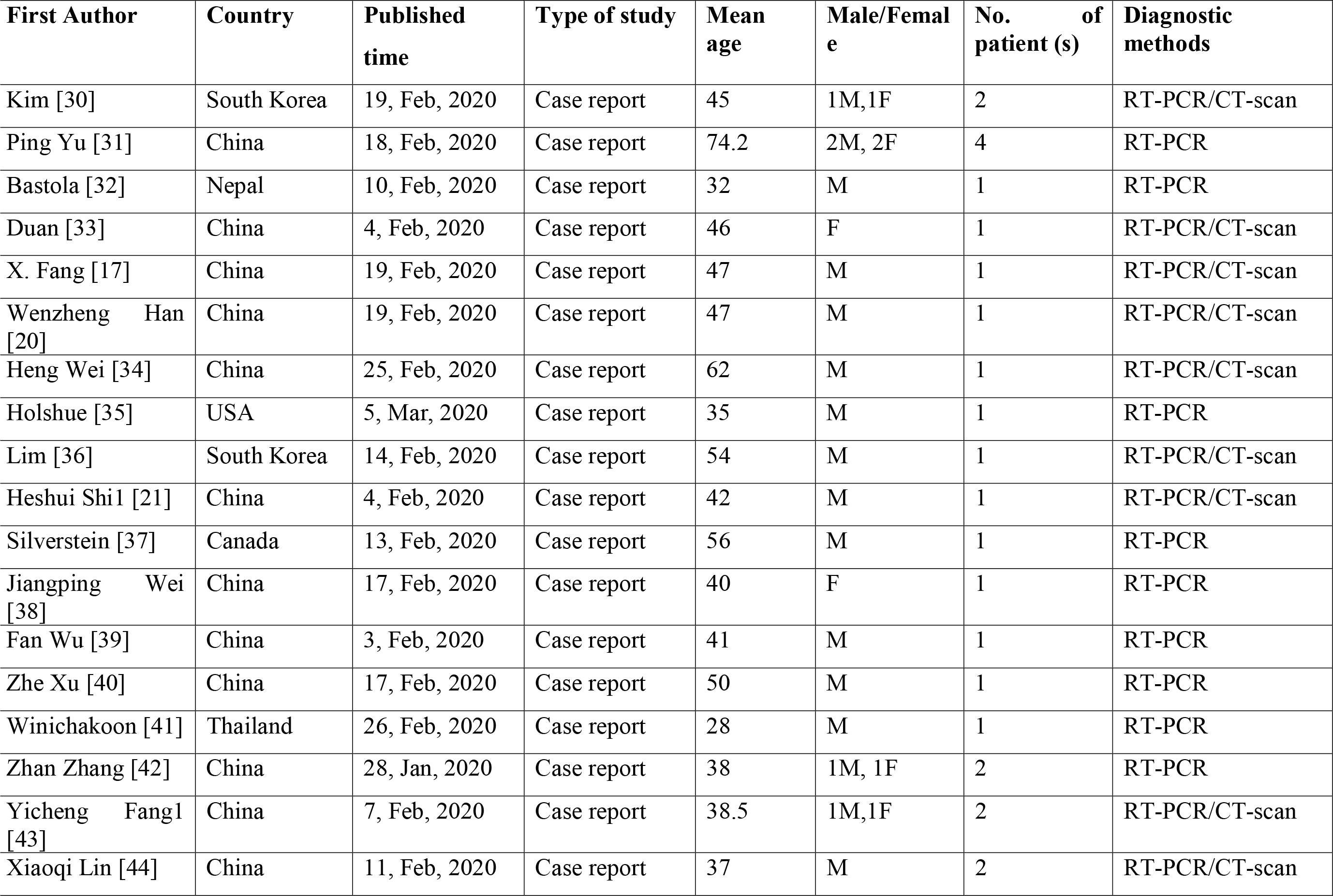
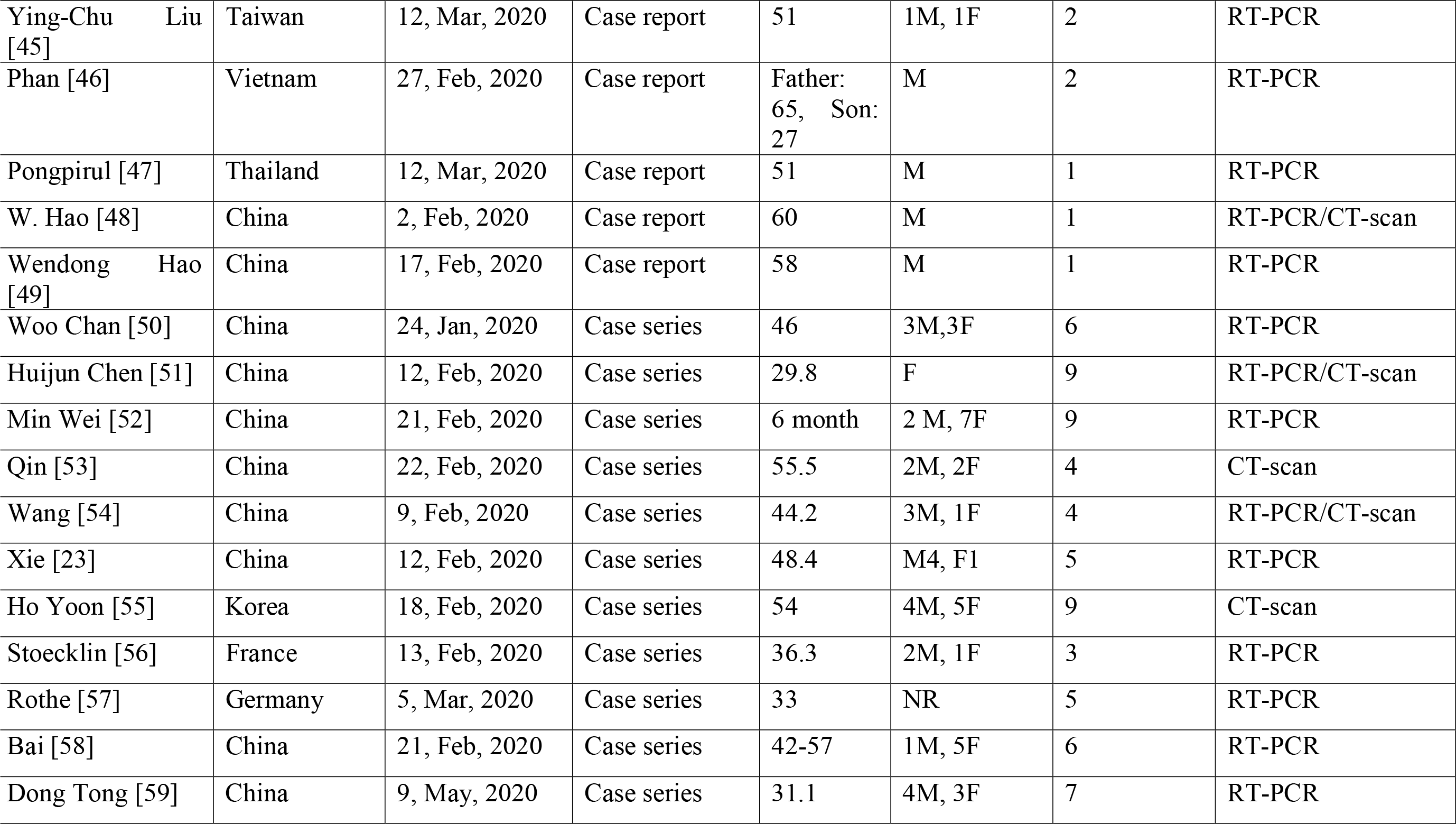
Characteristics of the included studies

**Table 2.**
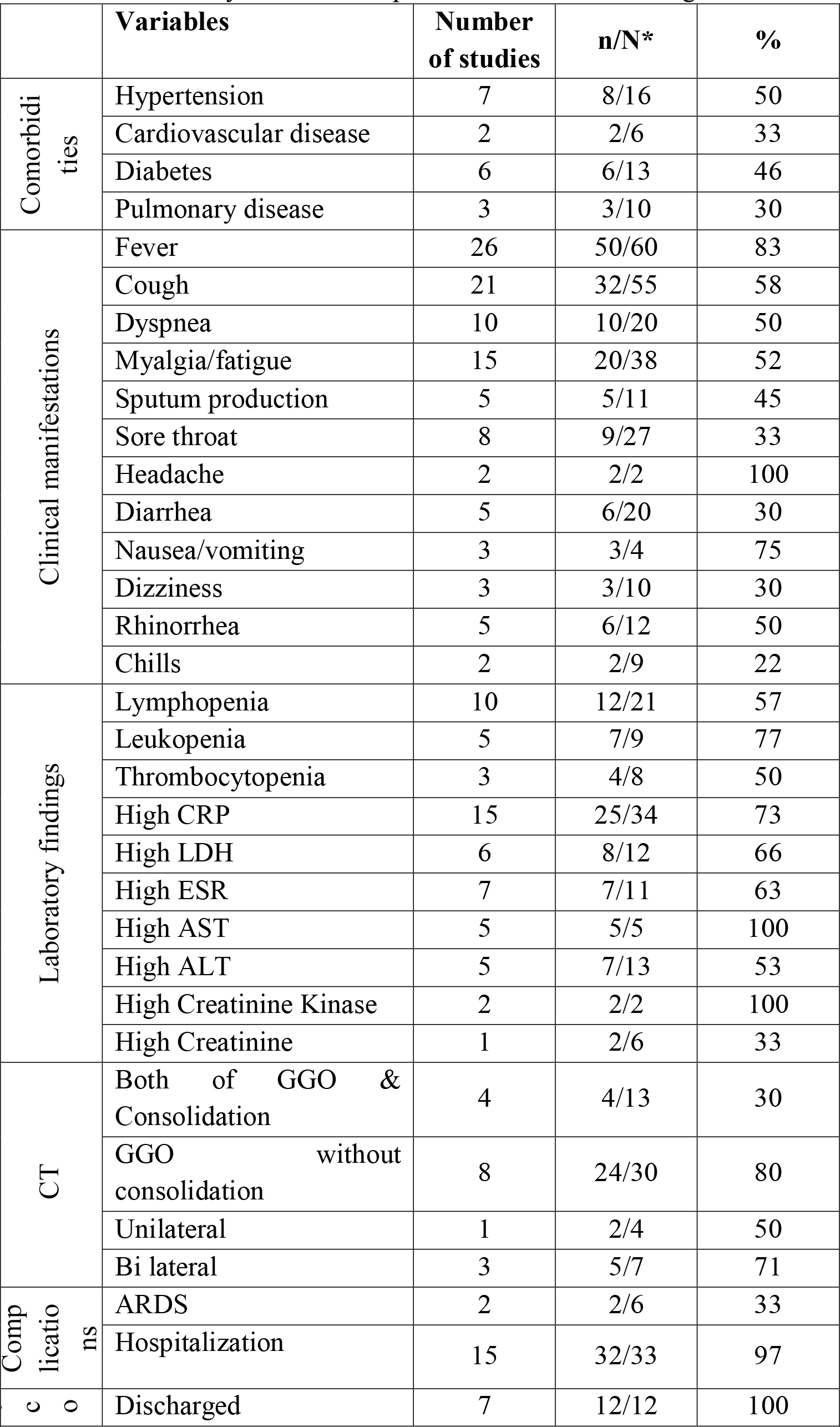
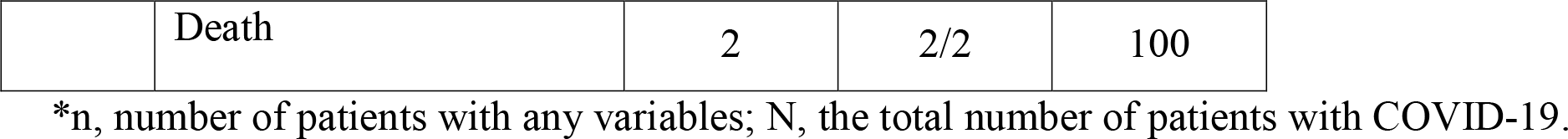
Summary of the case report and case series findings

|  | **Variables** | **Number**<br>**of studies** | **n/N\*** | **%** |
| --- | --- | --- | --- | --- |
| Comorbidi<br>ties | Hypertension | 7 | 8/16 | 50 |
|  | Cardiovascular disease | 2 | 2/6 | 33 |
|  | Diabetes | 6 | 6/13 | 46 |
|  | Pulmonary disease | 3 | 3/10 | 30 |
| Clinical manifestations | Fever | 26 | 50/60 | 83 |
|  | Cough | 21 | 32/55 | 58 |
|  | Dyspnea | 10 | 10/20 | 50 |
|  | Myalgia/fatigue | 15 | 20/38 | 52 |
|  | Sputum production | 5 | 5/11 | 45 |
|  | Sore throat | 8 | 9/27 | 33 |
|  | Headache | 2 | 2/2 | 100 |
|  | Diarrhea | 5 | 6/20 | 30 |
|  | Nausea/vomiting | 3 | 3/4 | 75 |
|  | Dizziness | 3 | 3/10 | 30 |
|  | Rhinorrhea | 5 | 6/12 | 50 |
|  | Chills | 2 | 2/9 | 22 |
| Laboratory findings | Lymphopenia | 10 | 12/21 | 57 |
|  | Leukopenia | 5 | 7/9 | 77 |
|  | Thrombocytopenia | 3 | 4/8 | 50 |
|  | High CRP | 15 | 25/34 | 73 |
|  | High LDH | 6 | 8/12 | 66 |
|  | High ESR | 7 | 7/11 | 63 |
|  | High AST | 5 | 5/5 | 100 |
|  | High ALT | 5 | 7/13 | 53 |
|  | High Creatinine Kinase | 2 | 2/2 | 100 |
|  | High Creatinine | 1 | 2/6 | 33 |
| CT | Both of GGO & Consolidation | 4 | 4/13 | 30 |
|  | GGO without consolidation | 8 | 24/30 | 80 |
|  | Unilateral | 1 | 2/4 | 50 |
|  | Bi lateral | 3 | 5/7 | 71 |
| Comp<br>licatio<br>ns | ARDS | 2 | 2/6 | 33 |
|  | Hospitalization | 15 | 32/33 | 97 |
| c<br>o | Discharged | 7 | 12/12 | 100 |
|  | Death | 2 | 2/2 | 100 |

\*n, number of patients with any variables; N, the total number of patients with COVID-19.

**Table 3.** Treatment agents used in the included studies

|  | Treatment | Agents | Number of studies | n/N* | % |
| --- | --- | --- | --- | --- | --- |
| Pharmacologic treatment | Antiviral Drugs | Lopinavir | 6 | 9/9 | 100 |
|  |  | Arbidol hydrochloride | 2 | 6/6 | 100 |
|  |  | Oseltamivir | 5 | 1/1 | 100 |
|  |  | Veletonavir | 1 | 1/1 | 100 |
|  |  | Remdesivir | 1 | 1/1 | 100 |
|  |  | Ribavirin | 1 | 1/1 | 100 |
|  |  | Ritonavir | 1 | 1/1 | 100 |
|  |  | Gancyclovir | 1 | 1/1 | 100 |
|  | Antibacterial Drugs | Moxifloxacin | 4 | 5/5 | 100 |
|  |  | Vancomycin | 1 | 1/1 | 100 |
|  |  | Cefepime | 1 | 1/1 | 100 |
|  |  | Meropenem | 2 | 2/2 | 100 |
|  |  | Piperacillin tazobactam | 2 | 2/2 | 100 |
|  |  | Sefoselis | 1 | 1/1 | 100 |
|  |  | Linezolid | 1 | 1/1 | 100 |
|  |  | Levofloxacin | 1 | 1/2 | 50 |
|  | Others | Methylprednisolone | 5 | 6/6 | 100 |
|  |  | Ambroxol Hydrochloride | 1 | 1/1 | 100 |
|  |  | Acetaminophen | 2 | 2/2 | 100 |
|  |  | Ibuprofen | 2 | 2/2 | 100 |
|  |  | Intravenous Immunoglobulin | 3 | 4/7 | 57 |
|  |  | Guaifenesin | 1 | 1/1 | 100 |
|  |  | Ondansetron | 1 | 1/1 | 100 |
|  |  | Interferon alpha-2b | 2 | 2/2 | 100 |
|  |  | Herbal patent medicine | 2 | 3/3 | 100 |
| Non-Pharmacologic treatment | Oxygen therapy | Non-invasive | 6 | 10/10 | 100 |

\*n, number of patients under treatment; N, the total number of patients with COVID-19.

**Figure 1.**
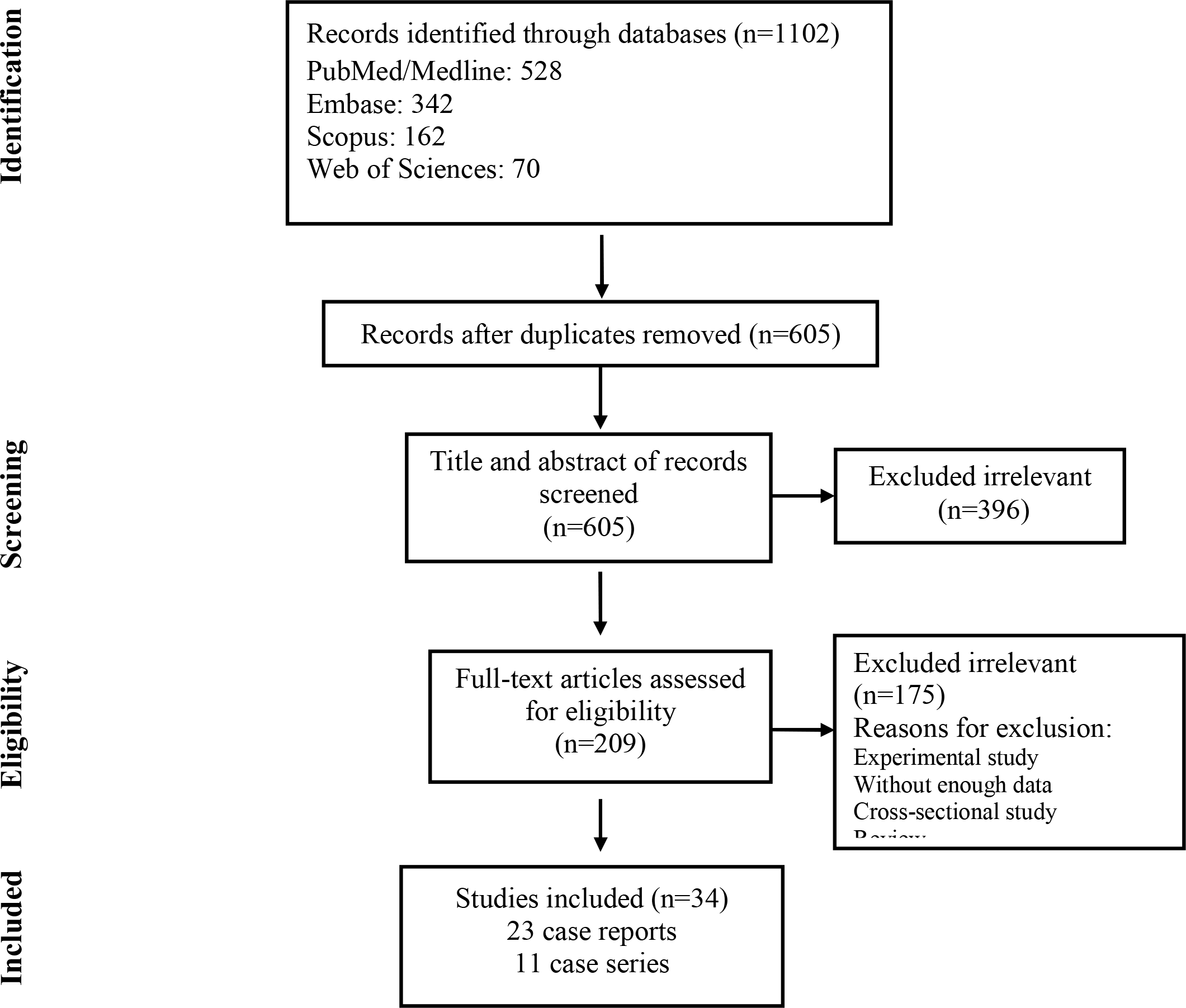
Flow chart of study selection for inclusion in the systematic review.

## Discussion

The 2019 novel coronavirus has been declared a public health emergency worldwide. The World Health Organization (WHO) declared COVID-19 a pandemic affecting 110 countries around the world with continued global spread. The 2019-nCoV is likely to be transmitted by asymptomatic individuals [7]. Asymptomatic transfer leads to lower prevalence estimates and higher transmission rates in the community. Until universal screening and vaccination become available, it is necessary to trace close contacts of those testing positive for COVID-19 and quarantining contacts to prevent asymptomatic transmission.

According to the articles we included, 2019-nCoV can only be transferred from person to person[8]. Chen et al suggested that they had no evidence of vertical transmission from mother to child [9]. Any person infected with 2019-nCoV can develop a clinical course of Covid-19. However, it is reported to cause the most severe symptoms such as respiratory failure in older men with comorbidities[10]. Children, teenagers, and younger people mostly showed a mild presentation of the disease [11].

Based on our reviewed articles, hypertension, diabetes, cardiovascular disease, and pulmonary disease were the most common morbidities among COVID-19 patients. This point was also mentioned in Alraddadi et al. study about MERS-CoV patients [12]. They showed that individuals with comorbidities like diabetes, smoking, and cardiovascular disease were associated with a more severe clinical course [12]. According to Yang et al., chronic diseases can debilitate the immune system and make proinflammatory conditions, leading to more severe infection and subsequently higher mortality rates [13].

According to the included studies, the most common clinical manifestations were fever, cough, dyspnea, and myalgia or fatigue. Less common clinical manifestations included nausea or vomiting, dizziness, rhinorrhea, and chills. Liu et al reported that infants had mild clinical manifestations and a better prognosis. Furthermore, some asymptomatic cases were seen among children.

The most common abnormal laboratory changes were lymphopenia, high concentrations of C-reactive protein, and elevated levels of aspartate aminotransferase; however, we do not know the exact pathogenesis and the reason for these alterations. Laboratory abnormalities may indicate the severity of disease and developing complications. According to Huang et al., most patients with secondary infection had a procalcitonin level greater than 0.5 ng/Ml and ICU patients had higher levels of prothrombin time and D-dimer [14]. Also, Liu et al. mentioned using hypoalbuminemia, lymphopenia, high concentrations of CRP, and elevated LDH to predict severity of acute lung injury[3]. Higher levels of angiotensin II are also proposed to be related to acute lung injury [3]. Meanwhile, non-survivors are suggested to have higher D-dimer and FDP levels, longer PT and aPTT, and lower fibrinogen and antithrombin levels [15].

CT scan as a diagnostic tool can be used to evaluate severity of pulmonary involvement and monitor clinical progression. CT scan has good sensitivity and can be used to establish COVID-19 diagnosis in patients who are highly suspicious based on epidemiologic history and clinical manifestations, but have negative PCR-based test results [16, 17]. It is important to highlight that the CT scan can be normal during initial days, and a normal CT scan in a suspected case would never definitely rule out the diagnosis of COVID-19 [18]. Moreover, the CT scan is dynamic in patients with COVID-19 and changes rapidly [19-21]. The earliest abnormal finding in the CT scan is the appearance of ground-glass opacities in peripheral and sub-pleural areas [22]. As the disease progresses, the GGO’s will expand and distribute more, most commonly to the right lower lung lobes. Later findings include consolidations, paving patterns, thickening of lobar fissures and adjacent pleura. Pleural effusion, hilar lymphadenopathies, and mediastinal lymphadenopathies are not common findings and have only been reported scarcely [23]. Lung pathology can progress to a “white lung” with low functional capacity or heal with some fibrotic remnants [24]. Dynamic changes in the lungs seen on CT imaging will continue even after the patient’s discharge [22]. In fact, CT scan findings have prognostic value in some patients, as Shi et al. have reported, deterioration on follow-up CT scan, old age, male sex, and underlying comorbidities are associated with poor prognosis.

ARDS was the most common complication among the confirmed COVID-19 patients; development of ARDS increased risk of patient morality[25]. Huang et al reported that the median time from onset of symptoms to the development of ARDS was 9 days [14]. Other complications were acute cardiac injury, acute kidney injury, secondary infection, and shock that lead to multiple organ failure [26, 27]. ICU patients in comparison to non-ICU patients were also more likely to have complications [28]. The mortality rate was higher in critically ill patients as well as in older patients with comorbidities and ARDS. Yang et al reported that the median duration from ICU admission to death was 7 days [25]. The window between the presentation to the time of ICU admission and/or development of ARDS is an optimal time for medical intervention.

There are many challenges in COVID-19 therapeutic strategies. There is currently no cure for COVID-19. However, pharmacologic and non-pharmacologic symptom management and supportive care measures should be given to all patients with symptomatic COVID-19. Other various therapeutic strategies have been trialed in patients with COVID-19 with the goal of slowing disease progression. There is a paucity of data surrounding the efficacy of treatments. Of the case controls and case series we included, antiviral agents including HIV protease inhibitors (lopinavir and ritonavir) as well as anti-influenza compounds (oseltamivir and arbidol) were used as treatment regimens. Unfortunately, we didn’t have enough information about the efficacy of each regimen; however, according to some studies, anti-HIV based medications could have benefits in more rapid improvement of clinical manifestations and decrease in viral load [19, 20, 29].

In conclusion, we discussed the clinical symptoms, laboratory abnormalities, common comorbidities, imaging modalities, and potential therapeutic options in COVID-19. We indicated that the most common symptoms were fever, cough, and dyspnea, but some young infected cases had no signs or symptoms. ARDS was the most common reported complication and was associated with poor prognosis. In the wake of the COVID-19 pandemic, countries are scrambling to produce enough RT-PCR diagnostic tests. Diagnostic information from other surrogate markers would be valuable if markers proved to be sensitive and specific. Namely, we learned that laboratory data like CRP may not only be related to the severity of disease, but it may be predictive of disease outcomes. Further studies are needed to relate quantified elevations in CRP to disease severity. Due to the high sensitivity of CT scan, it is considered as a good diagnostic tool. However, it should be kept in mind that a normal CT scan will never rule out the diagnosis of COVID-19 in a highly suspicious case based on history and clinical findings. Lastly, there are different therapeutic strategies for COVID-19 patients, but we don’t have enough data for their efficacy. Additional investigations including randomized controlled trials will be necessary to further our understanding in the treatment of COVID-19.

## Data Availability

All data are available

## Acknowledgment

This study was supported by Shahid Beheshti University of Medical Sciences, Tehran, Iran.

